# Rapid, point-of-care molecular diagnostics with Cas13

**DOI:** 10.1101/2020.12.14.20247874

**Authors:** Shreeya Agrawal, Alison Fanton, Sita S. Chandrasekaran, Bérénice Charrez, Arturo M. Escajeda, Sungmin Son, Roger Mcintosh, Abdul Bhuiya, María Díaz de León Derby, Neil A. Switz, Maxim Armstrong, Andrew R. Harris, Noam Prywes, Maria Lukarska, Scott B. Biering, Dylan C. J. Smock, Amanda Mok, Gavin J. Knott, Qi Dang, Erik Van Dis, Eli Dugan, Shin Kim, Tina Y. Liu, IGI Testing Consortium, Eva Harris, Sarah A. Stanley, Liana F. Lareau, Ming X. Tan, Daniel A. Fletcher, Jennifer A. Doudna, David F. Savage, Patrick D. Hsu

## Abstract

Rapid nucleic acid testing is a critical component of a robust infrastructure for increased disease surveillance. Here, we report a microfluidic platform for point-of-care, CRISPR-based molecular diagnostics. We first developed a nucleic acid test which pairs distinct mechanisms of DNA and RNA amplification optimized for high sensitivity and rapid kinetics, linked to Cas13 detection for specificity. We combined this workflow with an extraction-free sample lysis protocol using shelf-stable reagents that are widely available at low cost, and a multiplexed human gene control for calling negative test results. As a proof-of-concept, we demonstrate sensitivity down to 40 copies/μL of SARS-CoV-2 in unextracted saliva within 35 minutes, and validated the test on total RNA extracted from patient nasal swabs with a range of qPCR Ct values from 13-35. To enable sample-to-answer testing, we integrated this diagnostic reaction with a single-use, gravity-driven microfluidic cartridge followed by real-time fluorescent detection in a compact companion instrument. We envision this approach for Diagnostics with Coronavirus Enzymatic Reporting (DISCoVER) will incentivize frequent, fast, and easy testing.

## Introduction

Rapid, point-of-care nucleic acid detection is a critical component of a robust testing infrastructure for controlling disease transmission. Outbreaks such as the coronavirus disease 2019 (COVID-19) pandemic have highlighted limitations of the centralized diagnostic laboratory model and its lengthy sample-to-answer time. Laboratory-developed tests such as quantitative polymerase chain reaction (qPCR) are conducted in facilities that require labor-intensive personnel and equipment infrastructure for sample accessioning, nucleic acid extraction, thermocycling, and data analysis. Sample transport and result reporting time also greatly contribute to the turnaround times of centralized tests. The development of an easy to use microfluidic device paired with on-site detection would enable increased volume and access to rapid molecular testing.

To date, well over 2.5 million deaths from COVID-19 have resulted from over 115 million infections by its causative agent, severe acute respiratory syndrome coronavirus 2 (SARS-CoV-2). The myriad challenges of a high asymptomatic infection rate (Lavezzo et al. 2020), insufficient testing, and the narrow time window for molecular tests to be highly sensitive (Wölfel et al. 2020; Zhao et al. 2020) can be combated by broad deployment of on-site molecular diagnostics, such as upon entry into the workplace or classroom. There is also tremendous potential for community surveillance testing to augment clinical workflows, where positive cases are confirmed by referral to a more constrained supply of clinical-grade tests. Alternative sampling methods and test technologies can also help diversify the diagnostic supply chain, as the standard pipeline for clinical testing can be limited by RNA extraction kit or swab shortages (Vandenberg et al. 2020).

We therefore sought to develop a method that did not require RNA extraction or upper respiratory tract swabs and could be integrated into a rapid, automated microfluidic workflow. qPCR-based assays have been previously developed with direct sample spike-in, and typically use a high temperature for sample lysis (Bloom et al. 2020). Other approaches have exploited chaotropic agents, chemical reduction, and RNase inhibitors (Rabe and Cepko 2020; Myhrvold et al. 2018; Weickmann and Glitz 1982). Furthermore, saliva samples are reported to have 97% concordance with nasopharyngeal (NP) swabs (Iwasaki et al. 2020; Wyllie et al. 2020).

CRISPR-based detection is a promising new approach for nucleic acid diagnostics. These methods rely on the guide RNA-dependent activation of Cas13 or Cas12 nucleases to induce non-specific ssRNA or ssDNA nuclease activity, respectively, in order to cleave and release a caged reporter molecule (East-Seletsky et al. 2016; Gootenberg et al. 2017; Chen et al. 2018; Abudayyeh et al. 2016; Ramachandran et al. 2020; Ning et al. 2021). The released reporter can be quantitatively measured with a fluorescent detector to read out the test result. CRISPR-based detection is highly specific, but Cas13 nucleases alone can take up to two hours to reach attomolar sensitivity for diagnostic applications (Fozouni et al. 2020). In contrast, loop-mediated isothermal amplification (LAMP) performs highly sensitive nucleic acid amplification in under 20 minutes with attomolar limits of detection (LOD) (Nagamine, Hase, and Notomi 2002). However, despite the sensitivity and speed of LAMP, such isothermal methods are often prone to non-specific amplification (Hardinge and Murray 2019).

To address these challenges, we report DISCoVER (DIagnosticS with CoronaVirus Enzymatic Reporting), an RNA extraction-free test that combines two distinct amplification mechanisms for sensitivity with a Cas13-mediated probe for specificity (**Figure 1**). Following direct lysis and inactivation of saliva samples via denaturation and reduction, LAMP amplification is followed by T7 transcription to provide two layers of target amplification. To pioneer viral CRISPR testing in a sample-to-answer format, we integrated the DISCoVER workflow into a point-of-care microfluidic platform with a compact fluorescence reader for real-time detection.

**Figure 1.**
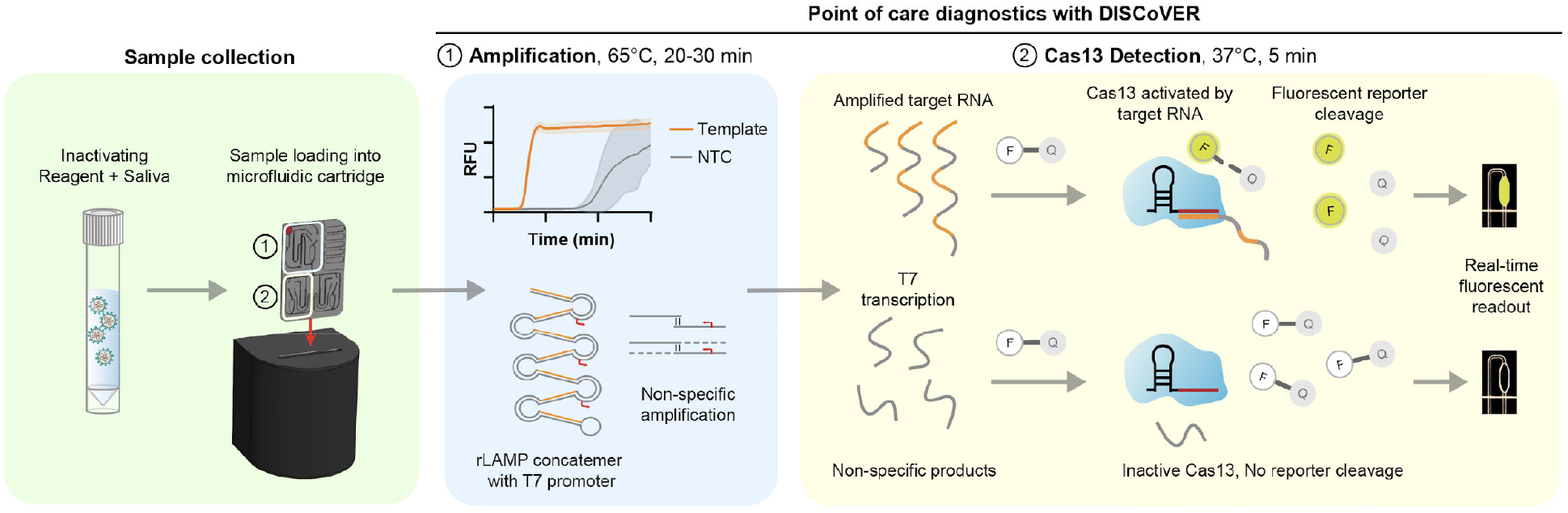
Schematic of point-of-care DISCoVER platform for molecular diagnostics. Patient samples, such as saliva, are collected and heat-inactivated in direct lysis buffer, followed by loading onto a single-use, gravity-driven microfluidic cartridge. The cartridge is then inserted into a companion instrument which automatically runs and the DISCoVER assay in a closed system to minimize reaction contamination. In Step 1, an initial rLAMP (RNA transcription following LAMP) reaction employs two mechanisms for amplification of target nucleic acids. Cas13 enzymes are programmed with a guide RNA to specifically recognize the desired RNA molecules over non-specifically amplified products. Subsequent activation of Cas13 ribonuclease activity, in Step (2), results in cleavage of reporter molecules for saturated signal within 5 minutes of CRISPR detection. In the left half of the cartridge, guide RNAs targeting SARS-CoV-2 enable rapid and selective detection of attomolar concentrations of virus. The mirrored half of the cartridge is used for an internal process control, enabling a negative test result by ensuring the presence of adequate patient sample. By exploiting template switching and CRISPR programmability, the point of care DISCoVER system can contribute to increased surveillance of diverse pathogens.

DISCoVER incorporates a 20-30 minute amplification step followed by Cas13 readout in under 5 minutes, and achieves attomolar sensitivity on unextracted saliva samples. We also multiplex the amplification with a process control based on detection of a human gene in order to confirm the presence of sufficient cellular material in the sample. We validated DISCoVER on a saliva-based sample matrix containing live SARS-CoV-2 virus and total RNA extracted from patient nasal swabs with Ct values ranging from 15-35. We observed a positive predictive value (PPV) of 100% and a negative predictive value (NPV) of 93.75% for the DISCoVER assay relative to qPCR. Additionally, contrived positive saliva and patient samples were assayed end to end on the microfluidic device followed by real-time fluorescent detection. The on-board process control enables discrimination between positive, negative, and invalid test results. Overall, the DISCoVER platform enables portable, rapid, and sensitive viral detection for point-of-care pathogen surveillance.

## Results

We first sought to compare the nucleic acid detection properties of Cas13 and Cas12 enzymes, assessing the reporter cleavage activity of *Leptotrichia buccalis* Cas13a (LbuCas13a) and *Lachnospiraceae* bacterium ND2006 Cas12a (LbCas12a) with matching guide RNA spacer sequences in a dilution series of their corresponding synthetic activator molecules (**Figure 2A**). LbuCas13a detection is significantly faster than LbCas12a at low activator concentrations - at 100 pM of activator, Cas13a reaches half-maximum fluorescence over 30 times faster than Cas12 (**Figure 2B**). However, the limit of detection of LbuCas13a is still in the femtomolar range after 60 minutes (East-Seletsky et al. 2017), which is already beyond the maximum sample-to-answer time that is likely to be relevant for a point-of-care assay.

**Figure 2.**
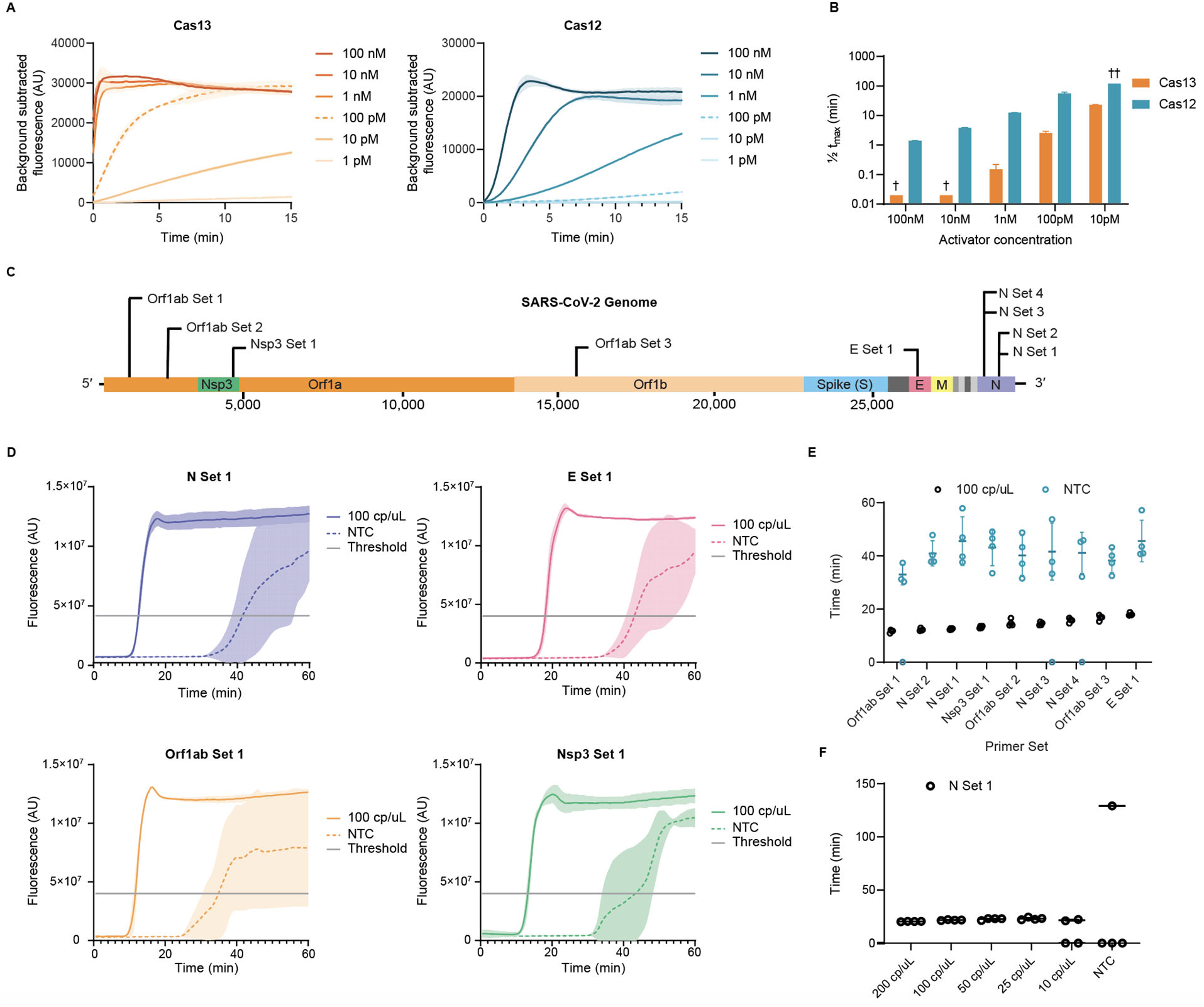
Direct nucleic acid detection with CRISPR-Cas enzymes and LAMP. **A**. Cas13 and Cas12 detection kinetics at varying activator concentrations. Values are mean ± SD with n = 3. **B**. Cas13 and Cas12 time to half-maximum fluorescence. †, Time to half maximum fluorescence was too rapid for reliable detection. ††, Time to half maximum fluorescence could not be determined within the 120 min assay runtime. Values are mean ± SD with n = 3. **C**. Schematic of SARS-CoV-2 genome sequence, with LAMP primer set locations indicated. **D**. Representative fluorescence plots of LAMP amplification of 100 copies/ µL of synthetic SARS-CoV-2 RNA or NTC. NTC, no-template control. Shaded regions denote mean ± SD with n = 3. **E**. Time-to-threshold of 9 screened LAMP primer sets, targeting synthetic SARS-CoV-2 RNA or NTC. Replicates that did not amplify are represented at 0 minutes. Error bars represent SD of amplified replicates. **F**. Limit of detection assay of LAMP using N Set 1 primer set. Replicates that did not amplify are represented at 0 minutes. Error bars represent SD of amplified technical replicates.

To achieve attomolar sensitivity within 30-40 minutes, we chose LAMP as a cost-effective and rapid method for isothermal amplification. LAMP employs a reverse transcriptase, a strand displacing DNA polymerase, and three sets of primer pairs to convert viral RNA to DNA substrates for LAMP. We screened nine LAMP primer sets targeting distinct regions across the length of the SARS-CoV-2 genome (**Figure 2C, Supplementary Table 1**). When targeted to SARS-CoV-2 genomic RNA fragments at 100 copies/µL, all sets resulted in positive LAMP signals. Maximum fluorescence was reached within 20 minutes for all primer sets, and time-to-threshold was determined via the single threshold Cq determination mode as indicated (**Figure 2D**). LAMP primer sets targeting Orf1ab Set 1, N Set 1, and N Set 2 consistently amplified in under 15 minutes (**Figure 2E**).

Each LAMP primer set resulted in a no-template control (NTC) signal, albeit with a delay relative to the positive condition containing viral RNA (**Figure 2D**). This high false-positive rate, potentially due to primer dimer formation, can in principle be reduced with a second probe that selectively recognizes the amplified nucleic acid sequence. We therefore sought to combine the sensitivity of LAMP detection with the specificity of Cas13 target recognition.

To avoid detection of non-specific amplification products by Cas13, the guide RNAs must have minimal sequence overlap with the primer sequences. Due to the complexity of LAMP concatemerization, LAMP primers are highly overlapping and amplify short target regions to increase the reaction speed (Notomi 2000). Our LAMP primer sets generated amplicons with primer non-overlapping sequences ranging from 1-60 nt. N Set 1, targeting the SARS-CoV-2 N gene, was chosen for further use due to its low time-to-threshold and an amplicon size capable of accommodating Cas13 guide RNAs (**Figure 2E, Supplementary Table 1**). We next performed a dilution series of genomic viral RNA and determined the N Set 1 LOD to be 25 copies/ µL (**Figure 2F**), comparable with previous studies that report LODs between 10-100 copies/ µL (Dao Thi et al. 2020; El-Tholoth, Bau, and Song 2020; Rabe and Cepko 2020).

Because Cas13 targets single-stranded RNA (**Figure 2A**), while LAMP amplifies DNA substrates, we employed transcription of the LAMP products to enable substrate compatibility. T7 RNA polymerase promoter sequences were incorporated into the LAMP primer sequences for subsequent transcription and Cas13 detection. We termed this conversion of LAMP amplification to RNA or rLAMP. LAMP employs three primer pairs: forward and backward outer primers (F3/B3) for initial target strand displacement, forward and backward inner primers (FIP/BIP) to form the core LAMP stem-loop structure, and forward and reverse loop primers (Floop/Bloop) for an additional layer of loop-based amplification (**Supplementary Figure 1**). Through multiple iterations of primer binding and extension, these stem-loop structures amplify into concatemers composed of inverted repeats of the target sequence (**Figure 3A**).

**Figure 3.**
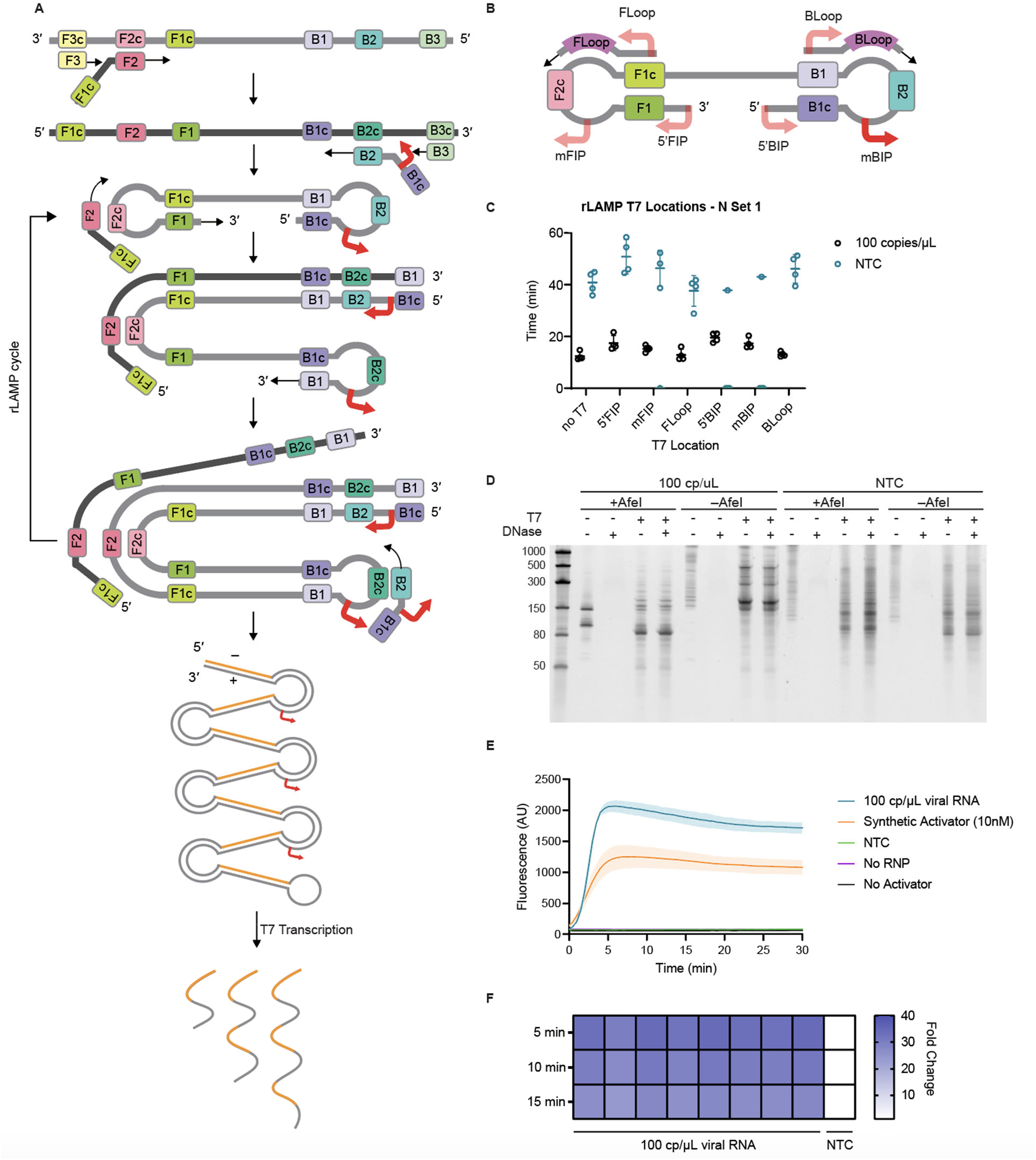
Development of RNA transcription following LAMP (rLAMP) for two layers of nucleic acid amplification. **A**. Schematic of rLAMP mechanism for exponential DNA amplification using F3/B3 and FIP/BIP primers, resulting in higher-order inverted repeat structures. Red arrows indicate location of T7 promoter sequence, inserted in the mBIP primer. Upon T7 transcription, the resulting RNA contains one or more copies of the Cas13 target sequence (orange). **B**. Schematic of the location of different T7 promoter locations on the rLAMP dumbbell structure and loop primers. **C**. rLAMP time to threshold of 6 distinct T7 promoter insertions. Replicates that did not amplify are represented at 0 minutes. Error bars represent SD of amplified technical replicates. **D**. Denaturing PAGE gels of mBIP rLAMP products to verify successful T7-mediated transcription. AfeI cleaves in the crRNA target region of templated products, which is expected to result in a single major transcribed species. **E**. Kinetics of T7 transcription and Cas13 detection on mBIP rLAMP products. RNP, ribonucleoprotein. Values are mean ± SD with n = 3. **F**. Cas13 detection of 8 technical replicates of mBIP rLAMP amplification on genomic RNA, depicted as fold-change over NTC at different reaction end-points.

To enable rLAMP, we systematically tested the insertion of T7 promoter sequences in three different regions of LAMP primers - on the 5’ end of the FIP and BIP primers (5’FIP/5’BIP), in the middle of the FIP and BIP primers (mFIP/mBIP), and on the 5’ end of the loop primers (FLoop/BLoop) (**Figure 3B**). Addition of the T7 promoter did not greatly affect rLAMP time-to-threshold of N Set 1, given viral genomic RNA at 100 copies/µL (**Figure 3C**). To confirm the target sequences were specifically amplified, we performed restriction enzyme digestion on the LAMP products using AfeI, which digests in the Cas13 guide RNA target region within the rLAMP amplicon (**Supplementary Figure 2A, 2B**). Lack of AfeI digestion in all NTC conditions confirmed that the NTC signal results from non-specific amplification of sequences lacking the guide-matching target sequence.

To test whether the T7 promoter was properly incorporated and functional in the rLAMP amplicon, we next performed *in vitro* transcription with T7 RNA polymerase. Denaturing PAGE analysis indicated that the viral template and NTC conditions resulted in significant RNA transcription for all primer sets (**Figure 3D**). As expected, AfeI digestion of the mBIP rLAMP product produced a single 147 nt product containing the T7 promoter (**Supplementary Figure 2A**). Subsequent T7 transcription resulted in the expected 85 nt RNA product (**Figure 3D**).

Next, we optimized reaction buffer conditions to support T7 transcription and Cas13 detection in a single step, followed by systematic screening of Cas13 cleavage activity in the presence of rLAMP products containing T7 promoter insertions in different rLAMP amplicons. We determined that the middle of the BIP primer (mBIP) insertion position resulted in the fastest detection and therefore chose this primer set for further studies (**Supplementary Figure 3**). Cas13 rapidly detects the rLAMP amplicon with a viral template, while avoiding detection of non-specific NTC amplicons, achieving over 10-fold change in signal over the NTC background in under two minutes (**Figure 3E**). Signal saturation occurred within five minutes, reaching a signal-to-background ratio of over 40.

To confirm replicability of the DISCoVER pipeline, we tested Cas13 detection on 8 replicates of mBIP rLAMP reactions with activator RNA included at a concentration of 100 copies/µL in a 20 µL reaction. All 8 replicates resulted in a > 25-fold increase in signal over NTC, which remains stable well beyond the 5 minute detection time employed here (**Figure 3F**).

With an amplification and detection protocol in place, we next optimized sample processing to establish a simple protocol of heat paired with chemical reagents to promote viral inactivation and dampen the activity of RNA-degrading nucleases present in saliva (Ostheim et al. 2020). We assayed two concentrations of the shelf-stable reducing agent TCEP (Tris(2-carboxyethyl)phosphine) paired with the ion chelator EDTA (ethylenediaminetetraacetic acid) (Rabe and Cepko 2020; Myhrvold et al. 2018), commercially available reagents such as QuickExtract buffers containing detergents and proteinases, and DNA/RNA Shield containing chaotropic guanidine thiocyanate.

To test the compatibility of these inactivating reagents with rLAMP, we created mock positive saliva samples by adding the reagents to heat-treated saliva at 75 °C (Chin et al. 2020) and adding SARS-CoV-2 genomic RNA at two different concentrations: 1000 cp/ µL and 200 cp/µL. In the absence of inactivating reagents, we were unable to detect any rLAMP signal, suggesting degradation of RNA in saliva by endogenous RNases present in the sample **(Figure 4A)**. In contrast, genomic RNA without saliva was rapidly amplified in under 15 min. Only the low concentration condition of TCEP-EDTA protected target RNA and preserved rLAMP sensitivity in all 4 replicates (**Figure 4A**). This reagent cocktail simultaneously breaks protein disulfide bonds and sequesters divalent cations. Its activity is expected to dampen RNase activity while simultaneously disrupting mucin gel formation, reducing variable saliva viscosity for simpler sample processing (Meldrum et al. 2018; Tabachnik, Blackburn, and Cerami 1981).

**Figure 4.**
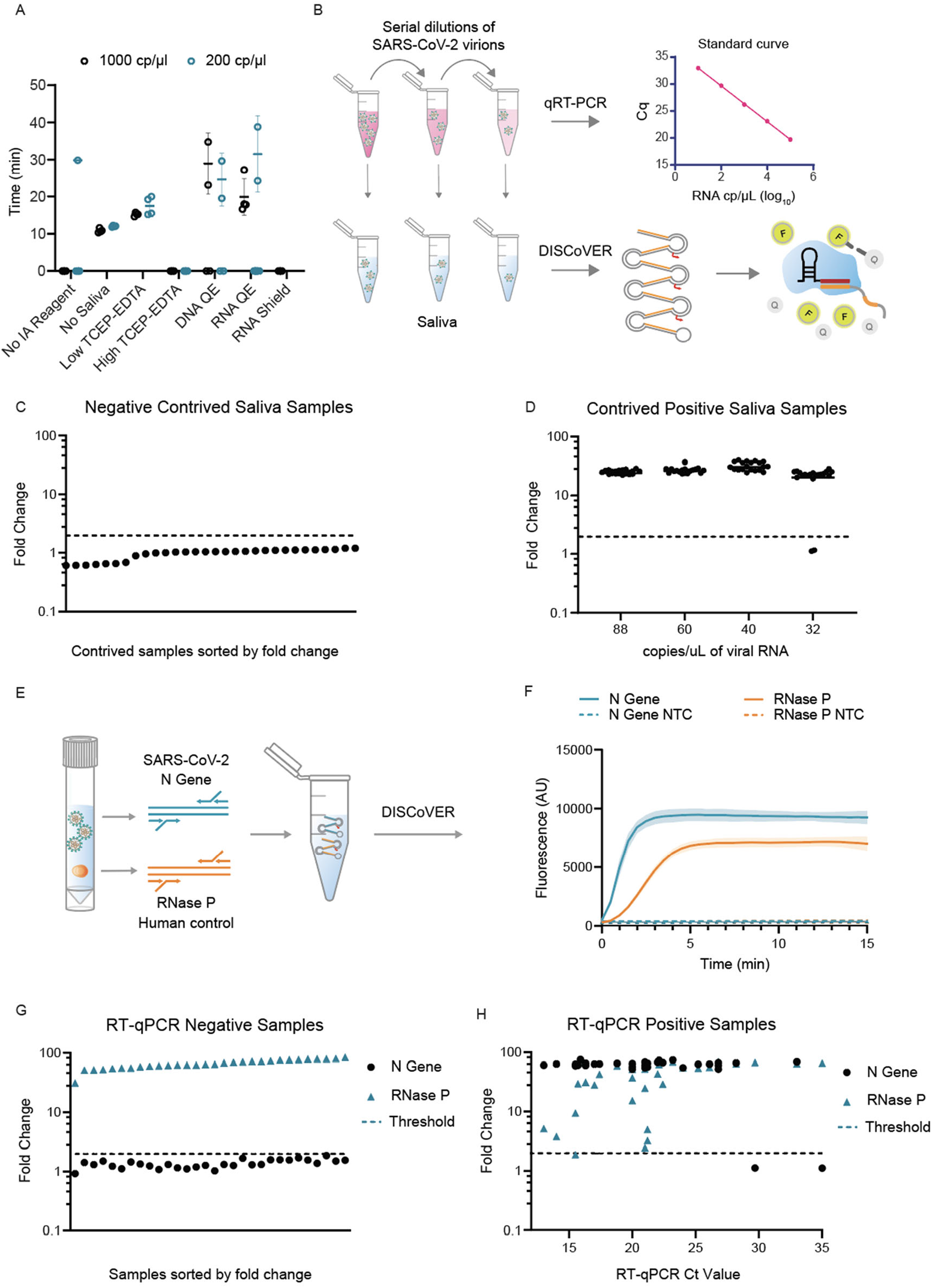
Development of DISCoVER for extraction-free detection of saliva and validation on patient samples. **A**. Direct saliva lysis conditions were tested for compatibility with the DISCoVER workflow. Replicates that did not amplify are represented at 0 minutes. Error bars represent SD of amplified technical replicates. IA: inactivation reagent; QE: QuickExtract. **B**. Schematic of contrived saliva sample generation, quantification via qRT-PCR, and detection via DISCoVER to determine analytical sensitivity. **C**. Fold-change in DISCoVER signal relative to NTC on SARS-CoV-2 positive saliva samples at 5 minutes of Cas13 detection. **D**. Fold-change in DISCoVER signal relative to NTC on 30 negative saliva samples collected before November 2019, at 5 minutes of Cas13 detection. **E**. Schematic of rLAMP multiplexing with SARS-CoV-2 (N Gene) and human internal control (RNase P) primer sets. **F**. DISCoVER signal of SARS-CoV-2 positive saliva samples after multiplexed rLAMP. Values are mean ± SD with n = 3. **G**. Fold-change in DISCoVER signal relative to NTC on 30 negative clinical nasal samples at 5 minutes of Cas13 detection. **H**. Fold-change in DISCoVER signal relative to NTC on 33 positive clinical nasal samples at 5 minutes of Cas13 detection.

We used the most recent guidelines provided by the FDA for Emergency Use Authorization (EUA) in May 2020 to determine the analytical sensitivity and specificity for DISCoVER on saliva samples (Food and Drug Administration, 2020). To determine the limit of detection, viral stocks were serially diluted in media and quantified with RT-qPCR relative to a standard curve generated from synthetic genomic RNA. These known concentrations of virus were spiked into negative saliva samples collected before November 2019 in BSL3 conditions and run through the DISCoVER workflow (**Figure 4B**). We performed 20 DISCoVER replicates for a range of virus concentrations, determining 40 cp/µL of virus in directly lysed saliva (**Figure 4C**) to be the lowest concentration tested where at least 19/20 replicates amplified successfully. To assay DISCoVER specificity, saliva samples from 30 different individuals negative for SARS-CoV-2 were tested without any false positive signal (**Figure 4D**). The addition of Influenza A H1N1 and H3N2, Influenza B, and human coronavirus OC43 synthetic RNA did not inhibit the detection of SARS-CoV-2, nor result in non-specific detection, further validating the specificity of this assay (**Supplementary Figure 4A**). Additionally, we validated DISCoVER on multiple SARS-CoV-2 variants of concern, including the highly transmissible B.1.1.7 variant (**Supplementary Figure 4B**) (Davies et al. 2021).

In point-of-care settings, successful sample inactivation and lysis would ideally be confirmed by an internal process control (**Figure 4E**). This is particularly important for successful saliva-based population screening, as some individuals can have high viscosity or mucin gel loads in their samples. According to FDA guidelines, a positive internal process control is necessary to determine if a sample is SARS-CoV-2 negative, indicating proper sample collection, nucleic acid extraction, assay set up, and reagent functioning (Catalog # 2019-nCoVEUA-01, CDC, 2019). If the internal process control is negative in a clinical specimen, the result should be considered invalid, unless SARS-CoV-2 is detected, in which case the sample is presumed positive. To achieve this, we multiplexed amplification by adding both N gene and human RNase P gene primers into the rLAMP step. Subsequent Cas13 detection of RNase P on saliva samples reached saturation within 5 minutes (**Figure 4F**).

Next, we validated DISCoVER in total RNA extracted from patient nasal swab samples. 30 negative and 33 positive patient samples were confirmed by qRT-PCR in a CLIA testing lab, and positive samples featured Ct values ranging from 13 - 35 (**Figure 4G, 4H**) (IGI Testing Consortium, 2020). We were able to correctly call 30 out of 30 negative samples within 5 minutes of Cas13 detection as RNase P was robustly detected while N gene signal remained below the threshold. We detected 31 out of 33 positive samples within the same timeframe, while also confirming RNase P signal for 32 out of 33 samples, indicating robustness of the workflow. In the single sample without RNase P detection, the clinical testing lab was also unable to detect RNase P, indicating potential sample degradation. Based on the clinical sample dataset, we report the sensitivity of DISCoVER to be 93.9% with 100% specificity. Additionally, we observe the positive predictive value (PPV) to be 100%, with a negative predictive value (NPV) of 93.75%.

Finally, we developed a compact and portable microfluidic device to perform the assay in a point-of-care setting. The chemistry was adapted to run on a gravity-driven microfluidic cartridge, with temperature-controlled reaction and detection chambers (**Figure 5A**). The device contains air displacement pumps for controlled fluidic flow, a resistive heater for the rLAMP chamber, and a thermoelectric cooler/heater (TEC) for Cas13 chamber cooling and heating (**Figure 5B**). The assay fluorescence detection was performed by a compact custom detector for real-time sample readout.

**Figure 5.**
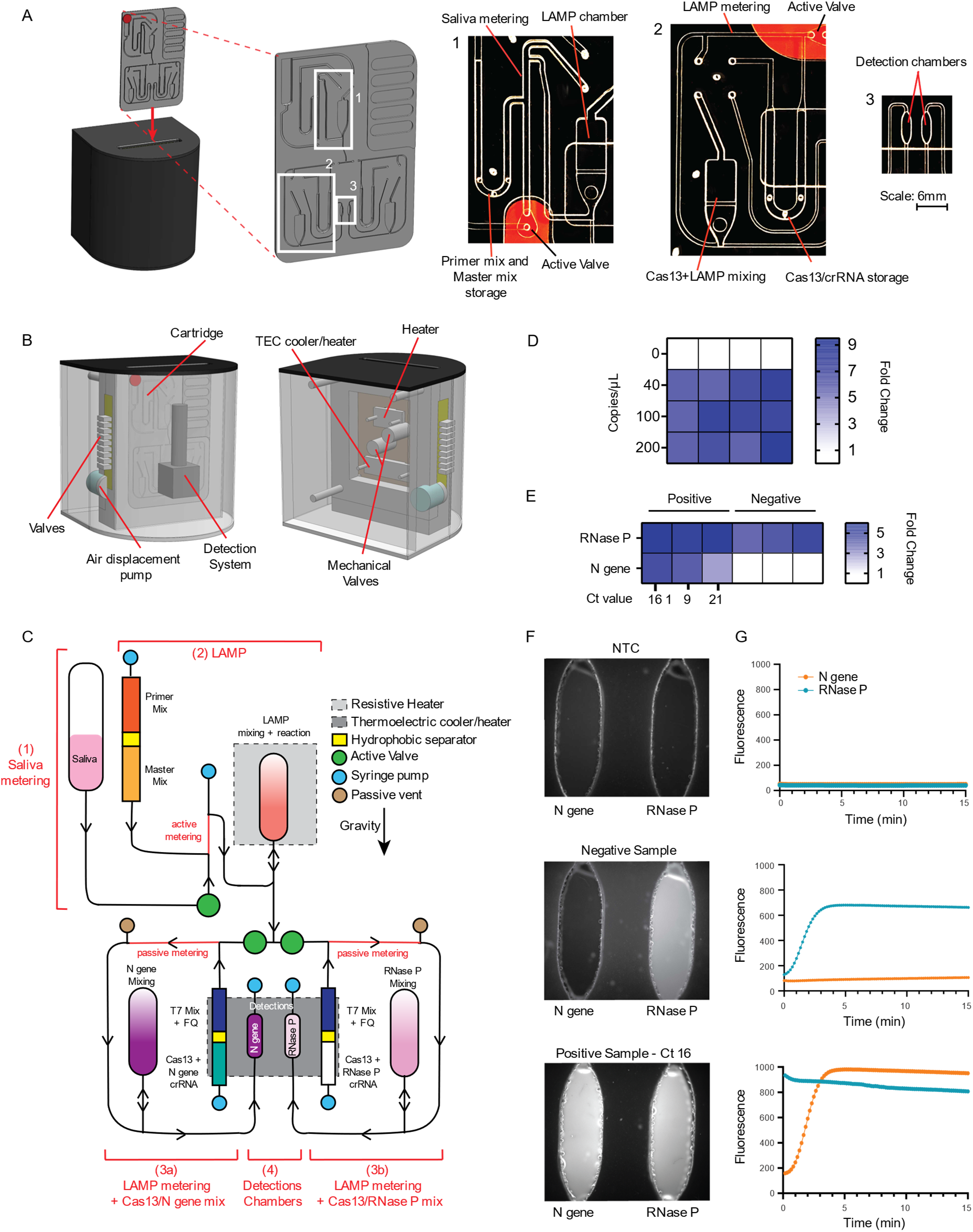
Implementation of a point of care, microfluidic-driven diagnostic platform. **(A)** Illustration of the final instrument in which the cartridge is inserted (left). Picture of the entire microfluidic gravity-driven cartridge (inset). On right, images of the sample metering and rLAMP reaction chamber (1), rLAMP metering and Cas13 mixing chamber (2), and the detection chamber (3). **(B)** Front (left) and rear (right) illustration of the instrument components, that include detection system, heaters, air-driven valves and mechanical valves. **(C)** Schematic of the cartridge function. The reaction can be separated in four steps: **1**. Saliva metering; **2**. amplification reaction; **3**. post-amplification metering + Cas13 mix; **4**. Cas13 reaction in detection chambers. Reagents stored on the cartridge are separated via a proprietary hydrophobic solution to avoid premature initiation of the reactions. After the LAMP reaction, the sample can be split into two reactions: The left part of the cartridge will expose the sample to N gene crRNA (**step 3a**) while the right side of the cartridge will act as internal control with only RNase P crRNA (**step 3b**). Saliva samples only went through **step 3a** while clinical samples went through **step 3a** and **step 3b. (D)** Heat map depicting the fold-change in DISCoVER signal on negative and positive saliva samples relative to the no-template control (NTC). **(E)** Heat map depicting the fold-change in SARS-CoV-2 RNA positive and negative clinical samples from nasal swabs, relative to NTC. **(F)** Fluorescent images of detection chamber for NTC cartridge (no viral sample), negative and positive clinical sample (with qRT-PCR Ct ranging from 16 to 21). The left detection chamber is specific for N gene detection whereas the right chamber is specific for RNase P. **(G)** Graph depicting raw fluorescence over time for both detection chambers (blue for RNase P and red for N gene) in NTC (left), negative (middle) and positive (right, Ct 16) samples.

A microfluidic cartridge was developed for positive or negative saliva sample detection (**Figure 5C**). Briefly, an inactivated positive saliva sample is loaded onto the cartridge and is actively metered into the rLAMP chamber, where it is mixed with N gene primers and rLAMP master mix. The solution is then heated to 65°C for 30 minutes while the Cas13 reagents are maintained at 25°C with the TEC. At the end of the rLAMP reaction, the active valve below the chamber opens, enabling gravity-assisted flow of 4 μL of the amplified solution into a metering channel. Only the left channel (**Figure 5C**, step 3a) was open, allowing for Cas13 reagents and its N gene-targeting crRNA to be pushed through the metering channel into the mixing chamber. The right side of the cartridge (**Figure 5C**, step 3b) was added in the later version of the cartridge. Cas13 mixed with the rLAMP solution was aspirated to the detection chamber and the reaction was monitored using a fluorescent detector. We successfully demonstrate detection of a dilution series from 200 - 40 cp/μl of SARS-CoV-2 genomic RNA in whole saliva, with a start-to-end reaction time of about 35 minutes (**Figure 5D**). This indicates that a point-of-care microfluidic platform for DISCoVER can successfully detect SARS-CoV-2 RNA from saliva samples with attomolar sensitivity.

Next, we optimized the cartridge design by multiplexing the detection of both N gene and RNase P during the rLAMP step to add an internal process control (step 3b in **Figure 5C**). Multiplexed rLAMP was performed before the amplified reaction was split into two Cas13 detection chambers, each containing a single crRNA targeting either the N Gene (left chamber, step 3a) or RNase P (right chamber, step 3b) rLAMP amplicon (**Figure 5C**).

We then tested qPCR-confirmed positive and negative clinical samples of total RNA from patient nasal swabs on this platform (IGI Testing Consortium, 2020). We tested the DISCoVER device on multiple clinical samples with Ct values ranging from 16 to 21 (**Figure 5E**), identifying RNase P signal for both SARS-CoV-2 negative (**Figure 5F and 5G**, middle) and positive (**Figure 5F and 5G**, right) samples. NTC cartridges without any viral sample were run initially to determine the baseline fluorescence, enabling us to determine the fold-change of a positive sample from baseline (**Figure 5F and 5G**, left).

## Discussion

Here, we report a sample-to-answer microfluidic platform for CRISPR-based molecular diagnostics. First, we developed an RNA extraction-free detection workflow, demonstrating the attomolar detection of SARS-CoV-2 RNA in unextracted saliva. DISCoVER’s combination of a 20-30 minute rLAMP step followed by T7 transcription and Cas13-based detection creates a rapid testing protocol with attomolar sensitivity and high specificity. As each modified LAMP product can serve as a substrate for transcription initiation, maximum Cas13 signal is reached in under five minutes due to rapid generation of nanomolar substrate concentrations (**Supplementary Figure 3**). The combination of sensitive nucleic acid amplification with CRISPR-mediated specificity and programmability has the potential to enable flexible diagnostics for diverse pathogen detection.

We demonstrated the DISCoVER system first on unextracted saliva, with the aim of enabling the first key steps for a point-of-care diagnostic. A saliva-based assay may not require medical personnel for sample collection, so is preferable to nasopharyngeal swabs, and the increased comfort of sample collection will likely incentivize patient compliance and commitment to frequent testing. It has also been shown that saliva is a reliable sample matrix for asymptomatic testing in community surveillance, as saliva samples have comparable viral titers to NP swabs (Wyllie et al. 2020). To diversify sampling supply chain, DISCoVER can also be applied to other samples such as NP swabs and self-administered nasal swabs.

In comparison with other CRISPR detection methods such as DETECTR and STOPCovid V2, DISCoVER does not employ a sample extraction or purification step (Broughton et al. 2020; Joung et al. 2020), eliminating reliance on commercial RNA extraction kits while maintaining comparable sensitivity. The direct lysis method employed here exploits common reagents for chemical reduction and ion chelation that are simple, widely available at low cost, and stable at room temperature. DISCoVER maintains attomolar sensitivity in comparison to other CRISPR based detection assays that include viral RNA extraction and purification (Broughton et al. 2020; Patchsung et al. 2020; Fozouni et al. 2020).

We further validate the ability of DISCoVER to detect multiple variants of SARS-CoV-2, specifically the B.1.1.7 variant of concern, which has been of broad medical interest due to its potential increased transmissibility (Davies et al. 2021). With additional development of mutant-specific DISCoVER probe sets, it may be possible to detect one variant of concern over another more selectively. Here, we optimized for pan-SARS-CoV-2 surveillance, using primers designed to detect as many SARS-CoV-2 variants as possible. To further test DISCoVER specificity, we assayed for cross-recognition of viral genomic RNA from other common respiratory pathogens, including influenza A H1N1 and H3N2, influenza B, and human coronavirus OC43. We did not observe a discernible effect on DISCoVER kinetics nor specificity.

We also demonstrate multiplexed SARS-CoV-2 detection with a human internal control during the DISCoVER amplification stage. This could be extended for multi-color detection using additional Cas enzymes with orthogonal cleavage motifs, each acting on specific reporters for distinct fluorescent channels (Gootenberg et al. 2018). Higher-order multiplexing can be further developed for influenza types A and B and other common respiratory viruses that would be desirable to detect in a single test. The inherent ability of the core enzymes in DISCoVER to convert between any RNA and DNA sequence implies that any pathogen that is inactivated and lysed by our protocol can be detected.

We validated DISCoVER on 33 positive and 30 negative patient samples, reporting a PPV of 100% and an NPV of 93.75% for total RNA extracted from nasal swabs. We observe 100% specificity and 93.9% sensitivity when compared with qPCR testing for samples with Ct values ranging from 13-35. Finally, we demonstrate the integration of DISCoVER with a point-of-care microfluidic platform and detection device. With simple heating and cooling elements and air displacement pumps, the gravity-driven microfluidic cartridge can perform the DISCoVER workflow on both contrived positive saliva and patient samples in 35 minutes with a real-time fluorescent readout. Further development and deployment of this point-of-care system will greatly facilitate frequent, on-site molecular diagnostics.

## Data Availability

All sequences used are presented in the manuscript or supplementary text. Any additional data is available upon request.

## Acknowledgments

We thank all members of the Hsu, Savage, Fletcher, and Doudna laboratories for support and advice, and Melanie Ott, Alexander J. Ehrenberg, Emeric Charles, and Brittney Thornton for helpful discussions. This work was supported by the Shurl & Kay Curci Foundation, anonymous donors, Emergent Ventures, NIH, and DARPA under award N66001-20-2-4033. The views, opinions and/or findings expressed are those of the authors and should not be interpreted as representing the official views or policies of the Department of Defense or the U.S. Government. We thank the National Institutes of Health for their support (P.D.H. R01GM131073, DP5OD021369, D.F.S. R01GM127463). J.A.D. is an Investigator of the Howard Hughes Medical Institute (HHMI). A.F. was supported by an NSF Graduate Research Fellowship.

## Author Contributions

As co-first authors, S.A., A.F. and S.S.C. designed and performed experiments, analyzed the data, and prepared the manuscript. P.D.H. conceived the project, designed experiments, provided overall supervision for this work, and wrote the manuscript with input from all authors. D.F.S. and J.A.D. provided experimental input, edited the manuscript, and co-supervised this work. B.C. provided microfluidic expertise and edited the manuscript. A.M.E., R.M., and M.X.T. designed and built the microfluidic cartridge and diagnostic instrument. S.S., A.B., N.S., M.A., A.H., and M.D.d.L.D. designed and built the optical instrument integrated with the microfluidic device, under the supervision of D.A.F. N.P. edited the manuscript and, with M.L., contributed to experimental design. Q.D. performed standard quantification experiments. D.C.S., T.Y.L., and G.J.K. purified proteins for assays and, with E.D. and S.K., provided biochemical expertise. A.M. developed computational methods for guide RNA selection with L.F.L., and S.B.B. and E.V.D. performed BSL-3 work with supervision from E.H. and S.S. The IGI Testing Consortium generated clinical samples.

