## Supplementary Text for "Rapid, point-of-care molecular diagnostics with Cas13"

**Supplementary Materials**

**Supplementary Table 1: LAMP primer sets screened, Related to Figure 2**

| Lamp Primer Set | Time to Amplification (min) | NTC-Amplification time delta (min) | Amplicon length (nt) | Reference |
|---|---|---|---|---|
| Orf1ab Set 1 | 11.6 | 21.3 | 9 | (Rabe and Cepko 2020) |
| N Set 2 | 12.2 | 28.7 | 26 | (Joung et al., n.d.) |
| N Set 1 | 12.5 | 33.1 | 46 | (Broughton et al. 2020) |
| Nsp3 Set 1 | 13.2 | 30.0 | 1 | (Park et al. 2020) |
| Orf1ab Set 2 | 14.8 | 25.4 | 1 | (Yu et al. 2020) |
| N Set 3 | 14.5 | 27.1 | 27 | (Zhang et al. 2020) |
| N Set 4 | 15.7 | 25.4 | 27 | (Zhang et al. 2020) |
| Orf1ab Set 3 | 16.7 | 21.5 | 0 | (Lamb et al., n.d.) |
| E Set 1 | 18.0 | 27.6 | 60 | (Broughton et al. 2020) |

**Supplementary Table 2: LAMP, rLAMP, qRT-PCR, and Cas13 sequences**

See attached file

**Supplementary Table 3: Fluidics calibration volumes**

| Volumes (µL) | Saliva | Primer Mix | Master Mix | Cas13+crRNA | T7mix + FQ |
|---|---|---|---|---|---|
| **Loaded** | 150 | 12 | 40 | 10 | 40 |
| **Lost** | - | 8 | 20 | 6 | 8 |
| **Reaction** | 16 | 4 | 20 | 4 | 32 |

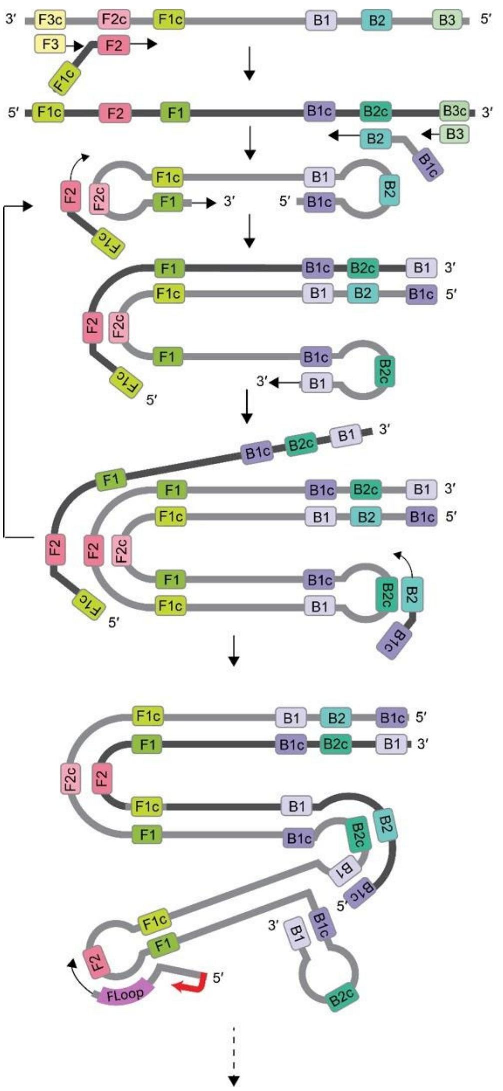

**Supplementary Figure 1**. **rLAMP with T7-Bloop primer mechanism.**

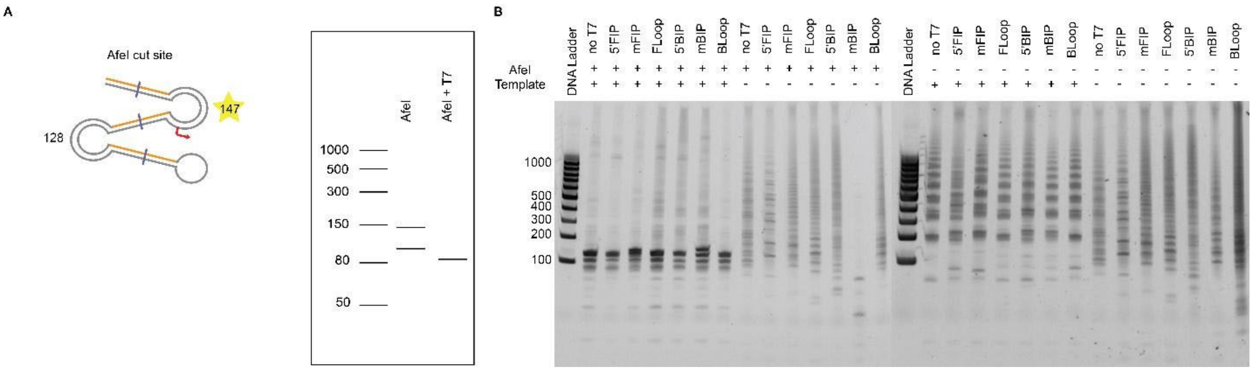


**Supplementary Figure 2. Validation of rLAMP products. A**. Schematic of primary mBIP rLAMP products and sizes upon AfeI digestion (left). Virtual gel depicting expected AfeI restriction digest bands (147 nt, 128 nt) and resulting transcription bands of mBIP rLAMP products. Only the 147 nt product is expected to contain the T7 promoter, and thus produce an ~85nt RNA product upon transcription. **B.** Restriction-digestion of each rLAMP product with each primer set containing the different T7 insertion positions.

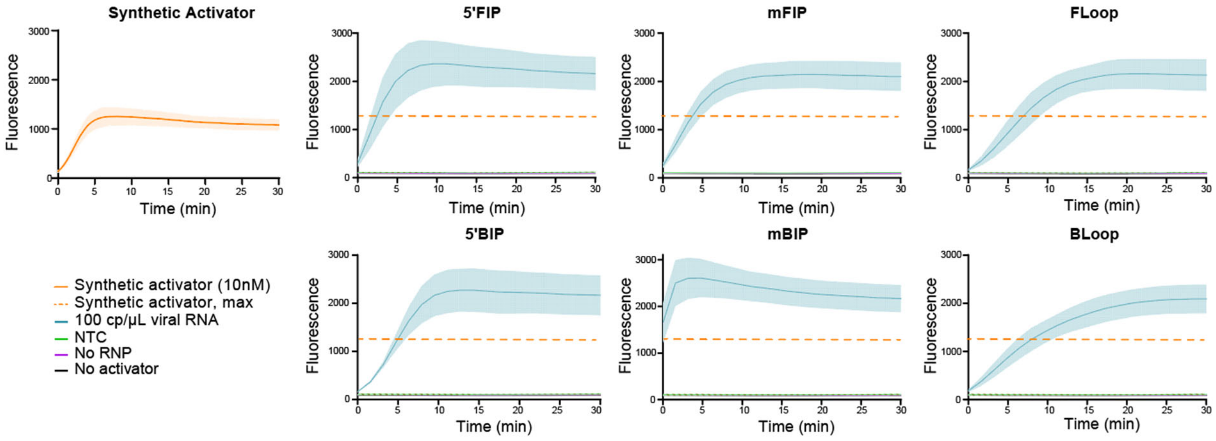


**Supplementary Figure 3**: Cas13 detection on all T7 insertion locations, with T7 transcription and Cas13 detection occurring in the same reaction. The maximum fluorescence of the synthetic activator at 10 nM is plotted (dashed line). Values are mean ± SD with n = 3. NTC, no template control. RNP, ribonucleoprotein.

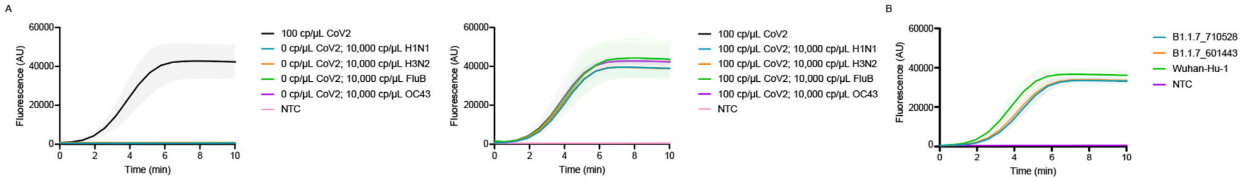


**Supplementary Figure 4. Specificity testing of DISCoVER Assay. A.** Detection of SARS-CoV-2 synthetic RNA in inactivated saliva in the presence of H1N2, H3N2, Influenza B, and human coronavirus OC43 synthetic viral RNA with N gene crRNA. Values are mean ± SD with n = 3. **B**. Detection of synthetic SARS-CoV-2 B.1.1.7. variants in saliva at 50 cp/μL with N gene crRNA. Values are mean ± SD with n = 3. NTC, no template control.

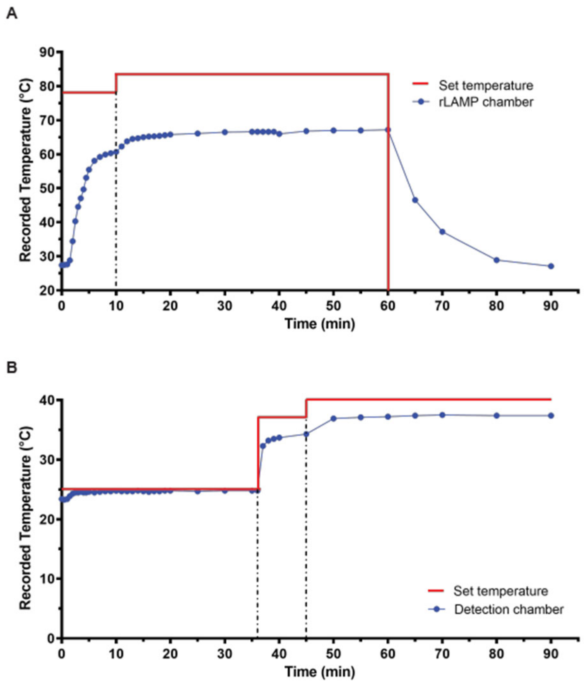


**Supplementary Figure 5. Temperature characterization of the cartridge using a thermal calibration cartridge.** Graph showing the temperature measured via integrated thermal-epoxy from the rLAMP chamber (**A**) and the detection chamber (**B**). On the x-axis, we showed the different timepoints at which the temperature was changed on the user interface.

### Methods

#### CRISPR-Cas direct nucleic acid detection

For *Lachnospiraceae* bacterium ND2006 Cas12a (LbCas12a), crRNA and the activatorDNA were synthesized via Integrated DNA Technologies (IDT). Detection with Cas12a was performed using 1X of NEB 2.1 buffer (B7202S), 100nM of LbCas12a protein, 100nM crRNA, varying concentration (100nM, 100nM, 10nM, 1nM, 100pM, 10pM) of dsDNA activator, and 200nM of Dnase Alert (IDT). LbCas12a and crRNA was pre-mixed at 10X concentration at a 1:1 molar ratio and incubated at room temperature (RT) for 15 minutes before adding it to the reaction. The reaction was heated at 37 °C with 15 sec interval detection with a Tecan Spark multimode microplate reader. For background subtraction, the fluorescence values of the no-activator reaction were subtracted from the fluorescence values of sample reactions at each time point. Maximum fluorescence was obtained by mixing DNase I and 200nM DNaseAlert with 1X NEB 2.1 buffer. This saturated maximum fluorescence value was divided in half; time to reach this half-maximum fluorescence was calculated for each concentration of activator.

For *Leptotrichia buccalis* Cas13a (LbuCas13a), the crRNA and the activator were commercially synthesized from Synthego. Detection with Cas13a was performed using 1X of Cas13 reaction buffer, 100nM of LbuCas13a protein, 100nM crRNA, varying concentrations (100nM, 100nM, 10nM, 1nM, 100pM, 10pM) of ssRNA, and 200nM of Cas13 reporter (IDT). LbuCas13a and crRNA was premixed at 10X concentration in 1:1 ratio and incubated at room temperature for 15 minutes before adding it to the reaction. The rest of the reaction, and time to half-maximum fluorescence, was performed or analyzed as described above. Maximum fluorescence was obtained by mixing RNase A and 200nM Cas13 reporter with 1X buffer. 5X Cas13 reaction buffer (pH 6.8) is composed of 100mM HEPES-Na pH 6.8 (Sigma), 250mM KCl (Sigma), 25mM MgCl2 (Sigma), and 25% glycerol (Thermo Fisher).

#### LbuCas13 protein production

The expression and purification of *Leptotrichia buccalis* Cas13a (LbuCas13a) was performed as previously described(East-Seletsky et al. 2017) with modifications, summarized here: $His_6$-MBP-TEV-tagged Cas13a was prepared in *E. coli* Rosetta2 (DE3) grown in TB at 37°C. At an $OD_{600}$ of 0.6–0.8, cultures were cooled on ice for 15 minutes prior to induction with 0.5 mM isopropyl β-D-1-thiogalactopyranoside (IPTG) and expression overnight at 16°C. Cell pellets measuring close to 10 mL in a 50 mL falcon tube were resuspended in 100 mL of Lysis buffer each. Soluble $His_6$-MBP-TEV Cas13a was clarified by centrifugation at 15,000 g, then loaded onto a 5 mL HiTrap NiNTA column (GE Healthcare) and eluted over a linear imidazole (0.01–0.3 M) gradient via FPLC (ÄKTA Pure). Following overnight dialysis and TEV digestion, LbuCas13a underwent purification by ion exchange and removal of MBP (5 mL HiTrap SP, MBP trap HP, GE Healthcare). Finally, size-exclusion chromatography was performed (S200 16/60, GE Healthcare) with 1 mM TCEP supplemented into the gel filtration buffer and peak fractions were pooled, concentrated and aliquoted into PCR strip tubes then snap frozen in liquid $nitrogen_2$.

**LbCas12a protein production**

The expression and purification of *Lachnospiraceae* bacterium ND2006 Cas12a (LbCas12a) was performed as previously described with the addition of 1mM EDTA in the dialysis buffer to prevent protein precipitation during His-tag removal by TEV protease (Knott et al. 2019).

**LAMP and rLAMP reactions**

LAMP and rLAMP DNA amplification reactions were conducted with 2X LAMP Mastermix (NEB, E1700), 0.4 µL LAMP dye (NEB, E1700), 2 µL of 10X primer mix (IDT; 16uM of FIP/BIP; 4uM Floop/Bloop; 2uM of F3/B3), and 7.6 µL of sample. 8 µL of sample was added in reactions in which no dye was added. For multiplexed LAMP, 1ul of RNase P primer with T7 promoter on the BIP 5' primer was added to 20ul reaction. Detection was performed at 65 °C for 30 - 60 min in a CFX96 Touch Real-Time PCR machine (Bio-Rad). Time to threshold was calculated using single threshold analysis mode. Limit of detection for LAMP was determined as the concentration where there was amplification observed with at least 95% of the samples. LAMP amplification reactions for clinical samples were conducted with 2X LAMP Mastermix, 2 µL of 10X N gene primer mix, 1ul of RNase P primer mix and 5 µL of sample in a 20ul reaction.

Twist Synthetic SARS-CoV-2 RNA Control 2 (Twist Bioscience, SKU 102024) was used as a template to perform the primer screen and establish LOD ranges. The volume of template to be used and the final concentration of template in the reaction was calculated based on the initial concentration provided by the vendor as 10^6 cp/µl.

**rLAMP assay development**

Restriction digestion of rLAMP DNA products was performed on 3 µL of amplified rLAMP product using 2 µL of AfeI (NEB R0652L), 2 µL 10X CutSmart Buffer, and 13 µL of water. This 20 µL of reaction was heated at 37 °C for 30 minutes. Undigested and digested rLAMP products were treated with 4 µL 1u/ µL RQ DNase I and 1 µL of 10X reaction buffer (Promega, M6101). The reaction was heated at 37 °C for 30 minutes, 1 µL of STOP solution (Promega M6101) was added, and incubated at 65 °C for 10 minutes. rLAMP products were analyzed using 15% TBE-Urea gels (Thermo Fisher, EC68855BOX).

**Direct saliva lysis**

SARS-CoV-2 negative saliva (Lee Biosolutions, 991-05-S-25) was used to check compatibility of saliva with the DISCoVER workflow. All saliva samples were collected prior to November 2019 as per vendor. When viral synthetic RNA was used, Twist Synthetic SARS-CoV-2 RNA Control 2 (Severe acute respiratory syndrome coronavirus 2 isolate Wuhan-Hu-1) was spiked in the commercial saliva and used as a template for mock samples. For limit-of-detection determination, SARS-CoV-2 viral seedstocks were spiked in negative saliva background under BSL-3 containment and used as a template for mock samples.

Commercial saliva was mixed with lysis reagents and heated at 75 °C for 30 minutes using a heat block. Twist synthetic RNA was added after completion of heat inactivation of saliva. Viral seed stocks were spiked in saliva in BSL3 facility prior to heat inactivation. Inactivating Reagent 1 was 2.5mM TCEP /1mM EDTA at 1X concentration when mixed with saliva. Inactivating Reagent 2 was 100mM TCEP/1mM EDTA at 1X concentration when mixed with saliva. Inactivating Reagent 3 was Quickextract DNA (Lucigen QE09050), Inactivating Reagent 4 was Quickextract RNA (Lucigen QER090150), and Inactivating Reagent 5 was RNA/DNAShield (Zymo 76020-420).

**Limit of detection (LOD) determination**

To determine the LOD, SARS-CoV-2(USA-WA1/2020; BEI Resources, NR-52281) viral stocks amplified once in Vero-E6 cells (ATCC) was diluted in media (DMEM + 10% FBS + 1% Penn/Strep) to obtain various concentrations, spiked into saliva, and inactivated by adding 1X of Low TCEP/EDTA inactivating reagent and heating the samples at 75 °C. The dilutions of virus in media were inactivated by heating at 75 °C for 30 minutes to comply with EH&S requirements (UC Berkeley) and RT-qPCR was performed using Luna® Universal Probe One-Step RT-qPCR Kit (NEB E3006) and E gene-targeting primers to determine the final concentration of viral RNA. Synthetic genomic RNA fragments (Twist Synthetic SARS-CoV-2 RNA Control 2) were used to obtain a standard curve for the calculations. The mock sample was detected as positive if the fold change (ratio of fluorescence value of sample to fluorescence value of no template control) is greater than 2 within 5 minutes. LOD for the mock samples was determined as the concentration where there was detection with at least 95% of the samples.

**Clinical sample RNA Extraction**

Clinical samples were obtained from the IGI Testing Laboratory, which utilizes a fully automated EUA workflow from Thermo Fisher (TaqPath COVID-19 Multiplex Diagnostic Solution) for both RNA extraction (MagMax Viral/Pathogen Nucleic Acid IsolationKit) and for RT-qPCR (TaqPath COVID-19 Combo kit) on clinical nasal swabs. These processes are detailed in (IGI Testing Consortium 2020).

**Validation with Clinical Samples**

We determined that a two fold change between a positive signal and negative control at five minutes into Cas13 detection is sufficient to distinguish mock positive and negative saliva samples. This experimentally derived threshold was used for calling positives and negatives in clinical samples. A sample is determined to be positive if both N gene and RNase P signal fold-change is above two-fold within 5 minutes. A sample is determined positive if N gene shows two-fold change within 5 minutes even if RNase P doesn't show two-fold change as it could indicate loss or degradation of human cellular material in saliva. A sample is determined to be negative if RNase P shows a fold change above 2 within 5 minutes while N gene signal does not appear. If both N gene and RNAse P fold change are below 2, the test is considered invalid.

**Cas13a guide design pipeline**

A LbuCas13a guide targeting SARS-CoV-2 was designed by prioritizing sensitivity, specificity, and predicted secondary structure. 26 candidate spacers of 20 nucleotides were identified within the N Set 1 amplicon region of the SARS-CoV-2 reference genome (wuhCor1), targeting both the forward and reverse strands. Sensitivity was determined by pairwise aligning available SARS-CoV-2 genomes from GISAID with the reference genome and calculating the number of mismatches between the sample and reference genome for each candidate spacer. The percentage of SARS-CoV-2 genomes detected by each spacer, with no more than one mismatch, was then calculated, and spacers matching fewer than 99% of SARS-CoV-2 genomes were discarded. To ensure specificity, candidate spacers that were determined to be sensitive were aligned to other reference genomes of human coronaviruses, specifically, SARS, MERS, 229E, NL63, OC43 and HKU1. Spacers that matched any of these other coronaviruses with two or fewer mismatches were discarded. The remaining spacers were also aligned to the human transcriptome, again allowing for two mismatches, and spacers with any alignments were discarded to ensure the guide would not be complementary to off-target human transcripts. To ensure that the spacer sequences would allow for Cas13a binding to the direct repeat scaffold, the concatenated repeat and spacer sequence was folded using RNAfold and evaluated for correct hairpin structure in the direct repeat and single strandedness in the spacer sequence. Out of 5 possible SARS-CoV-2 spacers passing our sensitivity, specificity, and structure criteria, one was chosen for use as a guide RNA. A control guide targeting RNase P was selected with the same criteria to avoid matching SARS-CoV-2 and human transcripts and to ensure proper RNA structure.

**T7 transcription**

Transcription was performed on amplified rLAMP product using 2 µL of 1mg/ml of T7 polymerase, 4 µL of 100mM NTP mix (NEB N0450), 1 µL of 200mM DTT, 4 µL of 5X transcription buffer, 2 µL of LAMP product and 7 µL of water. This 20 µL of reaction was heated at 37 °C for 30 minutes. 5X transcription buffer is composed of 150mM Tris-Cl, pH 8.1 at RT, 125mM $MgCl_2$, 0.05% Triton X-100 (Sigma Aldrich, X100), and 10mM spermidine and stored at -20°C.

**Cas13 detection**

Cas13 detection was performed as a 20 µL reaction using 2 µL of 1:100 diluted transcription product of LAMP, 1 µL 5X Cas13 reaction buffer, 2 µL of 2 µM Cas13 reporter, 2 µL of 10X RNP, 13 µL of 1X Cas13 reaction buffer. 10X RNP was made as a 2:1 ratio of Cas13:crRNA with final concentration of Cas13 as 20nM and incubated at RT for 15 minutes before adding it to the reaction. This 20 µL reaction was heated at 37 °C and read every 30s in TECAN Spark multimode microplate reader using FAM channel (Excitation:485; Emission:535).

**One-pot T7 transcription and Cas13 detection**

One pot T7 and Cas13 was performed by combining 2 µL of 1mg/ml of T7 polymerase, 20mM NTP mix, 10mM DTT, and 25mM $MgCl_2$ with the Cas13 reaction mix. 2 µL of 1:100 dilution of rLAMP product is used for 20 µL of one pot T7- Cas13 reaction. For testing clinical samples, 2ul of 1X rLAMP product is used for 20 µL of one pot T7- Cas13 reaction.

**DISCoVER specificity testing**

Synthetic viral RNA of Influenza A H1N1 (Twist Bioscience, SKU 103001), Influenza A H3N2 (Twist Bioscience, SKU 103002), Influenza B (Twist Bioscience, SKU 103003), and Human coronavirus OC32 (Twist Bioscience, SKU 103013) was spiked into inactivated saliva (Lee Biosolutions, 991-05-S-25), with and without Twist Synthetic SARS-CoV-2 RNA Control 2. rLAMP and one-pot T7 transcription as described above were performed with the N gene crRNA.

Synthetic viral RNA of SARS-CoV-2 variant B.1.1.7_710528 Control 14 (Twist Bioscience, SKU 103907) and SARS-CoV-2 variant B.1.1.7_601443 Control 15 (Twist Bioscience, SKU 103909) was spiked into inactivated saliva (Lee Biosolutions, 991-05-S-25). rLAMP and One-pot T7 transcription as described above were performed with the N gene crRNA.

**Instrumentation of point-of-care system**

A prototype instrument was designed and fabricated by Wainamics Inc. for an easy transition to a manufacturable end-use instrument. A combination of off-the-shelf components, custom-designed components (designed in SolidWorks), a custom electronic board based around a Teensy 3.2 microcontroller, and custom software (written in C) were integrated together to fabricate a prototype instrument, with user interface, controlling the functionality of the fluidic cartridge. The commercially available components used consisted of a Norgren Kloehn V6c syringe pump with 8-way rotary valve, Burkert Type 6724 3-way solenoid valves, Laird Thermal Systems temperature controllers, control fans (Delta Electronics, Inc), and resistive cartridge heaters (E3D). A custom graphical user interface on a Windows 10 PC allowed the user to control various functionality of the system and cartridge (i.e. actuate values, control volumes of air displaced into cartridge by pump, control flow rate of displaced air, control heater temperatures, etc). Script-recording allowed the user to generate and record instrument system scripts that were used to automatically run the assay on-board the cartridge to ensure precision and consistency across cartridge assay runs.

**Cartridge fabrication**

The cartridge was designed by Wainamics Inc. in SolidWorks. Prototype cartridges were designed and fabricated with manufacturability in mind. Laser-cut Polymethyl Methacrylate (PMMA) layers were laminated together using PCR-compatible pressure sensitive adhesives to achieve the desired channel geometries. All channel geometries were designed to be easily translated to, and replicated by, mass-manufacturing techniques such as injection molding and rotary die cutting. Each layer was cut by CO2 laser (BossLaser) and the entire cartridge was hand-assembled by Wainamics Inc. The PMMA material was sourced from Nitto Jushi Kogyo Co Ltd. The PSA material was sourced from Adhesive Research (AR90880 and AR 90712).

**Cartridge thermal calibration**

A cartridge containing thermocouples (Omega Engineering) embedded in thermal epoxy (Kona 870FT-LV-DP) was used for temperature calibration purposes. Power settings for resistive heaters and TEC were manually adjusted using the readings from the temperature calibration cartridge. A temperature calibration run using feedback from a thermal test cartridge is presented in **Supplementary Figure 5**. The difference between the set temperature and the recorded temperature increases at higher temperatures (not stable). It is therefore important to calibrate the system by verifying the set temperature necessary to match the required reaction temperature.

**Cartridge fluidics calibration**

Due to losses of reagents along the channels, multiple calibration experiments were performed to ensure correct reaction volumes (**Supplementary Table 3**). To measure the amount of volume
